# Drug library screen identifies inhibitors of toxic astrogliosis

**DOI:** 10.1101/2020.09.15.20195016

**Authors:** Ruturaj Masvekar, Peter Kosa, Christopher Barbour, Joshua Milstein, Bibiana Bielekova

**Author notes:** To whom correspondence should be addressed: Bibiana Bielekova, MD, Neuroimmunological Diseases Section (NDS), National Institute of Allergy and Infectious Diseases (NIAID), National Institutes of Health (NIH), Building 10, Room 5N248, 10 Center Drive, MSC1444, Bethesda, Maryland 20892, USA.

## Abstract

**Objective:** Multiple sclerosis is a chronic neuroinflammatory disorder, in which activated immune cells directly or indirectly induce demyelination and axonal degradation. Inflammatory stimuli also change the phenotype of astrocytes, making them neurotoxic. The resulting ‘toxic astrocyte’ phenotype has been observed in animal models of neuroinflammation and in multiple sclerosis lesions. Proteins secreted by toxic astrocytes are elevated in the cerebrospinal fluid of multiple sclerosis patients and reproducibly correlate with the rates of accumulation of neurological disability and brain atrophy. This suggests a pathogenic role for neurotoxic astrocytes in multiple sclerosis.

**Methods:** Here, we applied a commercially available library of small molecules that are either Food and Drug Administration-approved or in clinical development to an *in vitro* model of toxic astrogliosis to identify drugs and signaling pathways that inhibit inflammatory transformation of astrocytes to a neurotoxic phenotype.

**Results:** Inhibitors of three pathways related to the endoplasmic reticulum stress: 1) proteasome, 2) heat shock protein 90 and 3) mammalian target of rapamycin reproducibly decreased inflammation-induced conversion of astrocytes to toxic phenotype. Dantrolene, an anti-spasticity drug that inhibits calcium release through ryanodine receptors expressed in the endoplasmic reticulum of central nervous system cells, also exerted inhibitory effect at *in vivo* achievable concentrations. Finally, we established cerebrospinal fluid SERPINA3 as a relevant pharmacodynamic marker for inhibiting toxic astrocytes in clinical trials.

**Interpretation:** Drug library screening provides mechanistic insight into the generation of toxic astrocytes and identifies candidates for immediate proof-of-principle clinical trial(s).

## Introduction

Multiple sclerosis (MS) is a chronic inflammatory disease of the central nervous system (CNS), where activated immune cells directly or indirectly contribute to the loss of myelin sheath from CNS axons, leading to neurodegeneration. Affecting over 2.0 million individuals worldwide, MS is the most common non-traumatic neurological disorder in young adults (1).

Recruitment of immune cells from blood forms acute MS lesions in relapsing-remitting MS (RR-MS) (2,3). Development of focal lesions diminishes in later stages of MS (progressive MS [P-MS]), even though P-MS patients have levels of immune cell-specific cerebrospinal fluid (CSF) biomarkers indistinguishable from that of RR-MS (4), indicating persistent CNS inflammation. Consequently, the age-related decrease in the efficacy of immunomodulatory drugs on MS disability progression (5) has been attributed to inflammation compartmentalized to CNS tissue, largely inaccessible to current drugs. Alternatively, neurodegenerative mechanisms may drive CNS tissue destruction in P-MS. Inflammation-induced change in the phenotype and function of astrocytes towards “neurotoxic” (or “A1”) astrocytes identified in animal models of neuroinflammation represent a candidate neurodegenerative mechanism in MS (6-8).

Immunohistochemistry and *in situ* hybridization on post-mortem MS brain tissues showed the presence of A1 astrocytes in acute and chronic MS lesions (9).

Identification of CNS cell-specific protein clusters within the MS CSF proteome measured by a DNA-aptamer-based platform (i.e., Somascan®, Somalogic Boulder, CO, USA) detected only two clusters of proteins that reproducibly correlated with the rate of accumulation of disability and CNS tissue damage in the independent validation cohort of MS patients (10). One protein cluster was enriched for microglia-secreted proteins while the second cluster constituted proteins secreted mostly by inflammatory stimuli-activated astrocytes. Indeed, proteins of this cluster partially overlap with the “toxic astrocyte” signature identified by Liddelow et al. (9). Because this unbiased screen of CNS cell-enriched protein clusters supported a potential pathogenic role of toxic astrocytes in MS progression, we sought to develop an *in vitro* model of inflammation-induced neurotoxic astrocyte formation for a drug library screen, with two related aims: 1) to elucidate signaling pathways that underlie toxic astrocyte transformation; and 2) to identify Food and Drug Administration (FDA)-approved drug(s) with a reasonable toxicity profile for immediate use in proof-of-principle clinical trial of toxic astrocyte inhibition in MS.

## Methods

### Peripheral blood mononuclear cells (PBMCs) isolation and stimulation

PBMCs were isolated using density gradient centrifugation as described (10). PBMC (1×10^6^ cells/ml) were cultured in serum-free X-VIVO (Lonza), and either left untreated (unstimulated) or stimulated with lipopolysaccharide (LPS; Sigma-Aldrich, St. Louis, MO; 100 ng/ml) and CD3/CD28 microbeads (Invitrogen, Carlsbad, CA; at 1:1 beads to cells ratio) to activate simultaneously cells of innate and adaptive immunity. After 24 hours, cell-free supernatants were collected, aliquoted and stored at -80°C until further use. We validated that stimulated PBMC supernatants contained high levels of tumor necrosis factor α (TNFα) and interleukin 1 α (IL1α) using ELISA (R&D Systems, Minneapolis, MN; data not shown).

### Astrocyte cultures and treatments

Primary human astrocytes from cerebral cortex (ScienCell, Carlsbad, CA; Catalog# 1800; purchased on 03/2018, 06/2018 and 03/2019) were cultured (10^5^ cells/ml) as per manufacturer’s instructions. After 24 hours, cells were treated with 50% volume/volume of either unstimulated- or stimulated-PBMCs supernatants. 24 hours after treatment, cell-free culture supernatants were collected, aliquoted and stored at -80°C until further use. Cells were detached from the culture plate using trypsin-EDTA (Sigma-Aldrich, St. Louis, MO) for downstream applications.

### Immunostaining and flow cytometry

Astrocytes were immuno-stained for intracellular complement component 3 (C3) and serine protease inhibitor family A member 3 (SERPINA3) (9,11). Briefly, cells were resuspended in fixation/permeabilization solution (BD Cytofix/Cytoperm^TM^; BD Biosciences, San Jose, CA) for 20 min at 4°C, washed with permeabilization/wash buffer (BD Perm/WashTM; BD Biosciences) and stained with anti-C3 (Sigma-Aldrich; Catalog# GW20073F) or -SERPINA3 (R&D Systems; Catalog# MAB1295) antibodies conjugated with fluorescence-tag (Lightning-Link PE-Cy7 Antibody Labeling Kit; Novus Biologicals, Centennial, CO). Stained cells were washed twice and then analyzed using fluorescence-activated flow cytometry (BD LSR II Flow Cytometer, BD Biosciences, San Jose, CA).

### ELISA

C3 levels in astrocyte culture supernatants were measured using solid-phase sandwich ELISA (Genway Biotech, San Diego, CA; Catalog# GWB-1C0767). All samples were diluted 1:1 with bovine serum albumin (Sigma-Aldrich) and C3 concentrations were calculated using a standard curve according to the manufacturer’s instructions.

### Research subjects

All subjects were prospectively recruited under NIH IRB-approved protocol (Comprehensive Multimodal Analysis of Neuroimmunological Diseases of the Central Nervous System, ClinicalTrials.gov Identifier: NCT00794352), between 10/2008 and 04/2016. All subjects underwent neurological examination to derive the measures of neurological disability Expanded Disability Status Scale (EDSS) (12) and Combinatorial Weight-Adjusted Disability Scale (CombiWISE) (13). Composite magnetic resonance imaging (MRI) scale of CNS tissue destruction (COMRIS-CTD) was calculated from 3T brain MRI images as described (14). MS severity measures were calculated based on published algorithms either from EDSS: Multiple Sclerosis Severity Score (MSSS) (15) and Age-Related Multiple Sclerosis Severity (ARMSS) (16) or from CombiWISE: Multiple Sclerosis Disease Severity Scale (MSDSS) (5). MS diagnostic subgroups (RR-MS, secondary progressive MS [SP-MS] and primary progressive MS [PP-MS]) were classified using McDonald’s criteria, 2010 and 2017 revisions (17).

Healthy donors (HD) and untreated MS patients (Table 1 and Supplementary Table 1) were randomly divided into: training (n = 169) and validation cohort (n = 164), stratified on age, gender, MS type and MS severity (10).

**Table 1:** All subjects were divided into two cohorts, training and validation. Gender distribution (Chi-square test), age (ANOVA), and clinical outcome measures (EDSS, CombiWISE, COMRIS-CTD, MSSS, ARMSS and MSDSS; t-test) were compared across diagnostic categories (HD, RR-MS and P-MS [comprised of both SP-MS and PP-MS]).

| Diagnosis | Training Cohort (N = 169) |  |  |  | Validation Cohort (N = 164) |  |  |  |
| --- | --- | --- | --- | --- | --- | --- | --- | --- |
|  | HD | RR-MS | P-MS | P value | HD | RR-MS | P-MS | P value |
| **N (female/male)** | 8/10 | 35/23 | 44/49 | **0.2431** | 10/11 | 35/19 | 48/41 | **0.2965** |
| **Average** | 39.9 | 40.2 | 52.6 | **<0.0001** | 35.6 | 39.5 | 53.9 | **<0.0001** |
| **Age (SD)** | 15.6 | 11.3 | 10.4 |  | 11.0 | 10.6 | 9.0 |  |
| **range** | 19.4 - 70.3 | 18.0 - 68.7 | 22.0 - 65.8 |  | 19.7 - 57.4 | 18.3 - 67.9 | 29.8 - 74.7 |  |
| **Average** | - | 2.0 | 5.5 | **<0.0001** | - | 1.6 | 5.3 | **<0.0001** |
| **EDSS (SD)** | - | 1.5 | 1.6 |  | - | 1.0 | 1.5 |  |
| **range** | - | 0.0 - 6.5 | 1.5 - 8.5 |  | - | 0.0 - 6.0 | 2.0 - 7.5 |  |
| **Average** | - | 15.6 | 43.0 | **<0.0001** | - | 12.5 | 42.2 | **<0.0001** |
| **CombiWISE (SD)** | - | 10.8 | 14.7 |  | - | 6.9 | 13.6 |  |
| **range** | - | 2.2 - 50.4 | 9.4 - 84.5 |  | - | 2.4 - 35.5 | 14.5 - 70.8 |  |
| **Average** | - | 8.4 | 15.0 | **<0.0001** | - | 8.4 | 14.6 | **<0.0001** |
| **COMRIS-CTD (SD)** | - | 5.7 | 6.2 |  | - | 5.5 | 5.9 |  |
| **range** | - | 0.0 - 22.0 | 3.7 - 31.5 |  | - | 0.0 - 23.5 | 0.0 - 26.3 |  |
| **Average** | - | 4.0 | 6.8 | **<0.0001** | - | 3.6 | 6.8 | **<0.0001** |
| **MSSS (SD)** | - | 2.2 | 2.0 |  | - | 2.2 | 1.8 |  |
| **range** | - | 0.2 - 9.2 | 1.2 - 10.0 |  | - | 0.2 - 9.4 | 1.3 - 9.8 |  |
| **Average** | - | 3.5 | 6.5 | **<0.0001** | - | 3.1 | 6.2 | **<0.0001** |
| **ARMSSS (SD)** | - | 2.3 | 2.2 |  | - | 1.9 | 2.2 |  |
| **range** | - | 0.2 - 9.5 | 0.9 - 9.9 |  | - | 0.3 - 7.4 | 0.9 - 9.5 |  |
| **Average** | - | 1.4 | 2.1 | **<0.0001** | - | 1.3 | 2.1 | **<0.0001** |
| **MSDSS (SD)** | - | 0.7 | 1.1 |  | - | 0.5 | 0.9 |  |
| **range** | - | 0.4 - 3.5 | 0.3 - 5.3 |  | - | 0.5 - 2.6 | 0.7 - 4.4 |  |

### CSF collection and processing

CSF were collected by lumbar puncture and processed as per standard operating procedure (18). CSF aliquots were prospectively labeled using alphanumeric code, and immediately stored on ice after collection. Cell-free CSF supernatants were collected by centrifugation (335gx10 minutes at 4°C), aliquoted, and stored at -80°C. Personnel processing CSF were blinded to diagnoses and clinical outcomes.

### DNA-aptamer-based multiplex proteomics

CSF and cell culture supernatants were analyzed using the Slow Off-rate Modified Aptamers scan (SOMAscan; SomaLogic Inc., Boulder, CO) (19). CSF supernatants were analyzed using the 1.1K SOMAscan platform (analyzes 1128 proteins, available from June 2012 to October 2016), and cell culture supernatants were analyzed using the 1.3K platform (analyzes 1317 proteins, available from October 2016).

### Neurotoxicity

The human neuroblastoma cell line, SK-N-SH (ATCC® HTB-11^TM^; ATCC, Manassas, VA), was cultured according to the manufacturer’s instruction. After 24 hours, cells were treated with either unstimulated- or stimulated-astrocyte culture supernatants (50% v/v). After 24 hours apoptotic neurons were analyzed using Annexin V-FITC (TACS® Annexin V Kit; Trevigen Inc., Gaithersburg, MD) as described (20).

### Drug library screening

Astrocytes were plated (10^5^ cells/ml) on poly-L-lysine (Sigma-Aldrich)-coated 96-well cellculture plates (100 µl/well). After 24 hours, cells were treated with either unstimulated- or stimulated-PBMCs supernatants (50% v/v) in the presence of a respective drug (10 µM or 100 nM, 1431 drugs; Selleckchem LLC, Houston, TX; Catalog# L1300) or dimethyl sulfoxide (DMSO, a drug solvent; Sigma-Aldrich; control). 24 hours after treatment, supernatants were collected, and C3 levels were analyzed using ELISA. Percent change in absolute C3 secretion for a drug treatment with respect to control was calculated: ([C3 secretion by stimulated astrocytes with a drug treatment – C3 secretion by unstimulated astrocytes] / [C3 secretion by stimulated astrocytes with DMSO treatment – C3 secretion by unstimulated astrocytes]) * 100. Cytotoxic effects of respective drug treatments were analyzed using the 3-(4,5-dimethylthiazol-2-yl)-2,5-diphenyltetrazolium bromide (MTT) assay (ThermoFisher Scientific, Waltham, MA) as per manufacturer’s instructions.

### Statistical analyses

To differentiate biomarkers specific for MS biology from physiological age- and gender-differences, CSF SOMAscan values for all subjects were adjusted for age and gender dependency within HD subgroup as described (21).

Group-wise comparisons were performed using Analysis of Variance (ANOVA). When statistically significant (p < 0.05) differences were found, pairwise multiple comparisons using Tukey’s p-value adjustments were performed. Kruskal-Wallis ANOVA examined differences between diagnostic subgroups in biomarker values within the training cohort. When statistically significant differences were found, pairwise multiple comparisons using Dunn’s p-value adjustment were performed. Only statistically significant (adjusted p-value < 0.05) differences were then validated in an independent validation cohort.

Relationship between astrocyte-specific biomarkers and clinical outcomes were examined using Spearman correlations. Only statistically significant (p < 0.05) correlation coefficients in the training cohort were then validated in an independent validation cohort.

Drugs from the library screen were grouped based on their known targets (153 groups). Mean C3 concentrations (normalized to control treatment) of drugs that did not induce substantial toxicity (i.e., MTT > 75%), at 100 nM concentrations, were calculated and compared with C3 levels of control (i.e. 100%) using ANOVA. P-values were adjusted for multiple comparisons using Benjamini and Hochberg False Discovery Rate (FDR).

## Results

### Expression of toxic astrogliosis biomarkers

Induction of toxic astrogliosis by stimulated PBMCs was verified by demonstrating upregulation of intracellular C3 and SERPINA3, known markers of toxic/reactive astrocytes (9-11,22)(Figure 1A) and by observing that toxic-astrocyte-conditioned medium (50% v/v) induces apoptosis of neuronal cell line SK-N-SH (Figure 1B).

**Figure 1:**
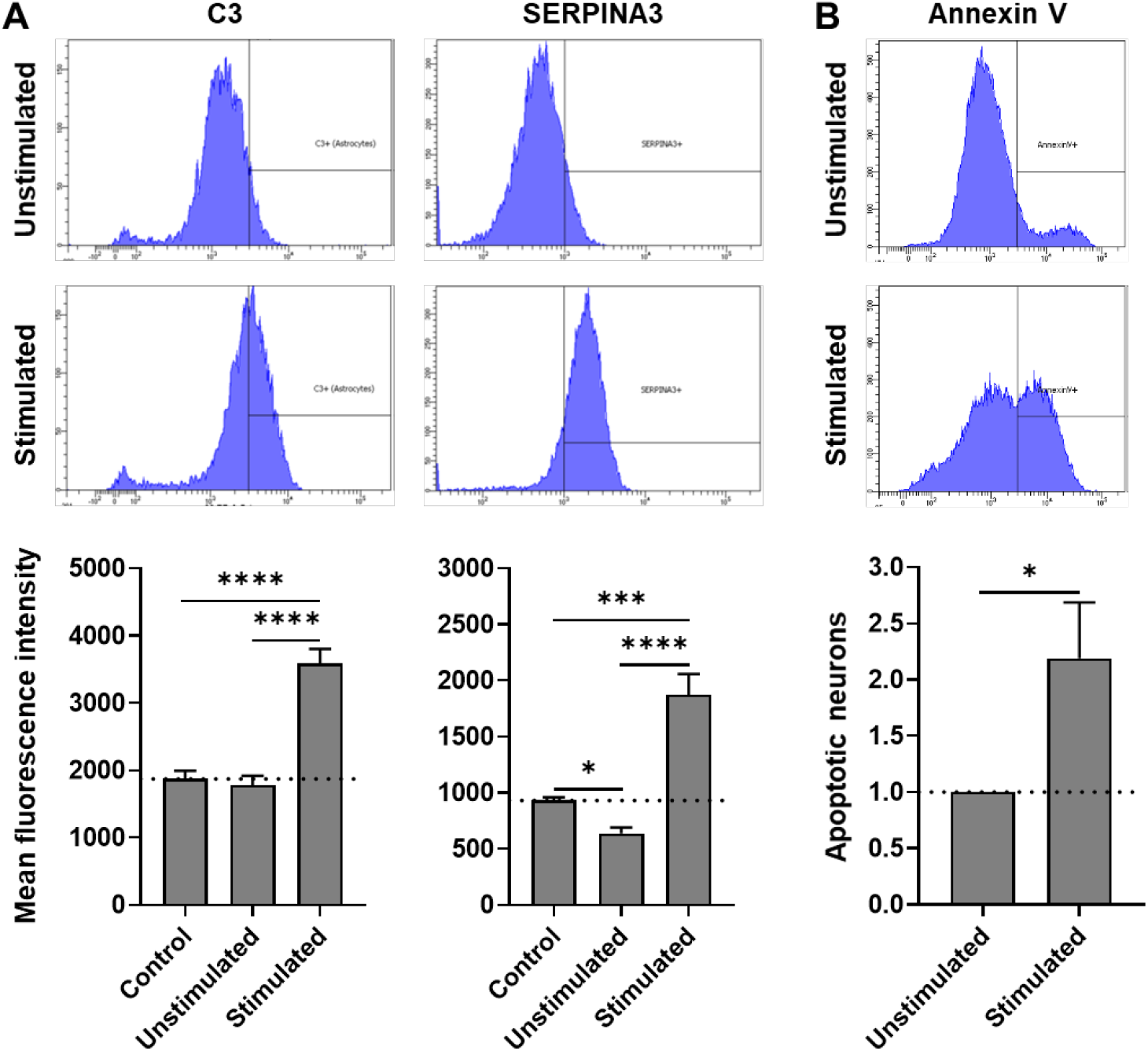
(A) Representative flow cytometry images of astrocytes immuno-stained for C3 and SERPINA3, 24 hours after treatment with unstimulated- or stimulated-PBMCs supernatants. Normalized, intracellular expression (mean fluorescence intensity) of C3 and SERPINA3 under different culture conditions is represented in respective charts. Expression across treatment groups (n = 3) were compared using ANOVA (Tukey’s multiple comparisons test); *p < 0.05, ***p < 0.0005 and ****p < 0.0001. (B) Representative flow cytometry images of neuroblastoma cell line (SK-N-SH) stained with Annexin V after treatment with unstimulated- or stimulated-astrocyte conditioned media (Astrocyte supernatant) for 24 hours. Normalized % of apoptotic (Annexin V+) neurons, compared (n = 4) using paired t test; *p < 0.05.

### Identification of inflammation-induced, astrocyte-secreted biomarkers

Cell culture supernatants from unstimulated- and stimulated-astrocytes were collected before (0 h) and after (24 h) the respective treatments and analyzed using DNA aptamer-based proteomic assay. Stimulation index for each protein was calculated by the ratio of relative fluorescence units (RFU) under stimulated (24h / 0h) versus unstimulated (24h / 0h) conditions (Supplementary Table 2). We arbitrarily defined measured proteins as inflammation-induced, astrocyte-secreted biomarkers if their stimulation index was > 5.

18 proteins were identified as inflammation-induced, astrocyte-secreted biomarkers: C-X-C motif chemokine ligand (CXCL-6, 9, 10 and 11), matrix metalloproteinases (MMP-3, 10, 12 and 13), C-C motif chemokine ligand (CCL-7, 8 and 20), SERPINA3, C-X3-C motif chemokine ligand 1 (CX3CL1), TNFα induced protein 6 (TNFAIP6), C3, complement factors (CFB and CFH) and interleukin 1 receptor like 1 (IL1RL1).

### Analysis of biomarkers across disease diagnosis subgroups

Age- and gender-adjusted SOMAscan values for inflammation-induced astrocyte-secreted biomarkers were compared among diagnostic subgroups (HD, RR-MS, P-MS [comprised of both SP-MS and PP-MS]) in the training cohort. Statistically significant differences were validated in an independent validation cohort. CXCL10 and C3 were significantly elevated only in RR-MS patients compared to HD, and MMP13 was elevated only in P-MS patients. While, SERPINA3 was significantly elevated in both MS subgroups (RR-MS and P-MS) (Figure 2A and Supplementary Table 3).

**Figure 2:**
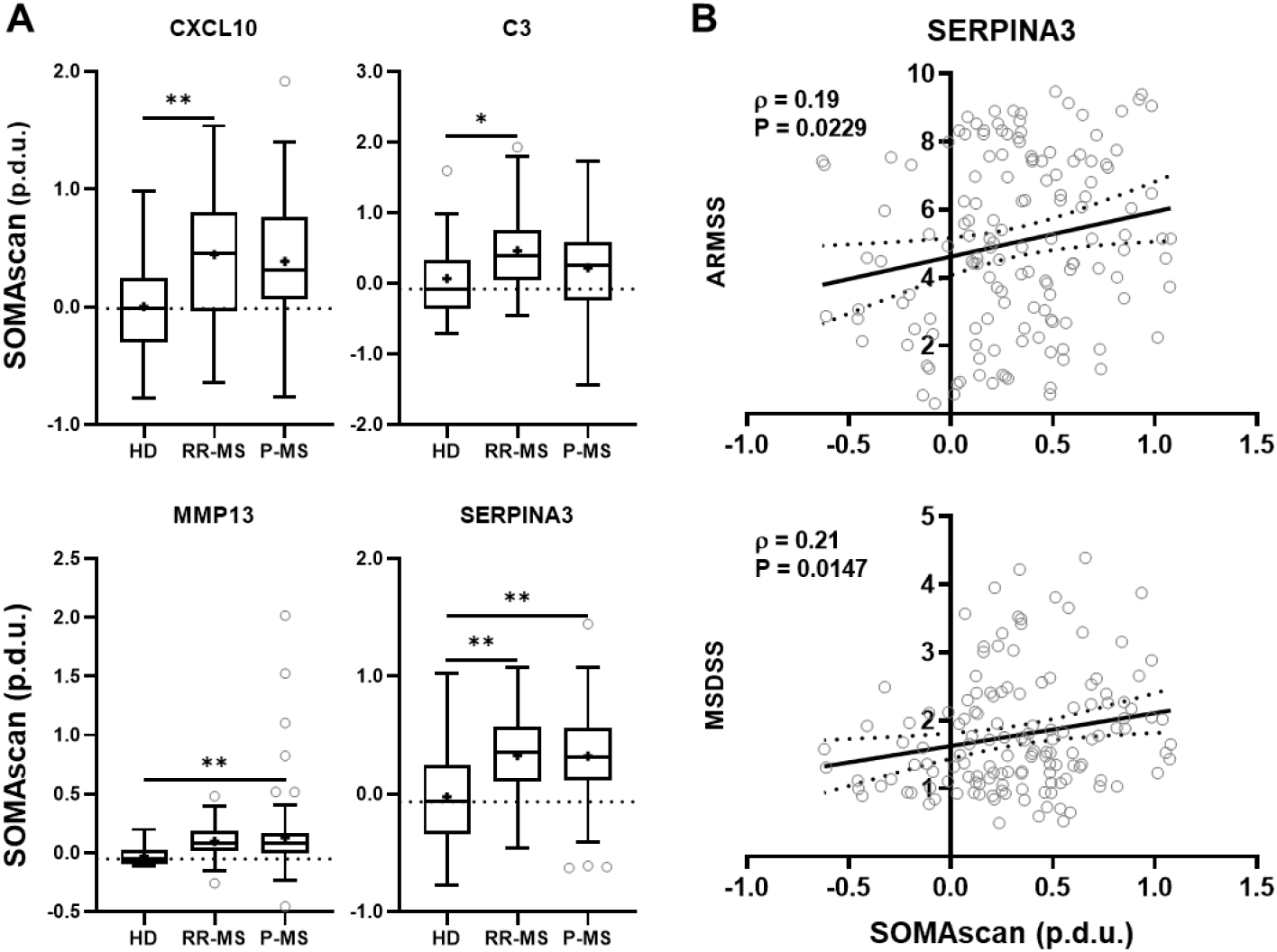
(A) Age- and gender-adjusted SOMAscan values (procedure defined unit, p.d.u.) of inflammation-induced, astrocyte-secreted biomarkers were compared across disease diagnostic subgroups using Kruskal-Wallis ANOVA (Dunn’s multiple comparisons test). *p < 0.05 and **p < 0.005. The ‘+’ sign represents mean of respective subgroup and dotted line indicates the median of the HD subgroup. (B) Correlations between SOMAscan values and clinical outcome measures were analyzed using Spearman analyses. The solid line indicates the best-fit line for linear regression between respective variables and the dotted line represents the 95% confidence interval. Spearman’s rank correlation coefficient (ρ) and adjusted P values are represented on respective correlation plots. Only statistically significant and reproducible results from the validation cohort are presented.

### Correlation analysis between biomarkers and clinical outcome measures

To determine how inflammation-induced, astrocyte-secreted biomarkers change with MS progression, RFU values for inflammation-induced, astrocyte-secreted biomarkers were correlated with the disability outcomes EDSS (12), CombiWISE (13), and with MRI scale of CNS tissue destruction (COMRIS-CTD) (14). No statistically significant correlations were observed (Supplementary Table 4).

To assess the potential pathogenic role of toxic astrocyte-secreted biomarkers in MS, we correlated biomarker RFUs (adjusted for natural aging and gender effects as described in Methods) with validated measures of MS severity: MSSS (15), ARMSS (16), and MSDSS (5). Only SERPINA3 had reproducible correlations with two MS severity outcomes, ARMSS (ρ = 0.19 and adjusted p = 0.0229) and MSDSS (ρ = 0.21 and adjusted p = 0.0147) (Figure 2B) in the independent validation cohort. This suggests that CSF SERPINA3 levels reflect the pathogenic role of toxic astrocytes in MS-associated CNS tissue destruction.

### Drug library screening

We applied a commercially available drug library to *in vitro* model of inflammation-induced astrocytes to identify therapeutic targets to impede induction of the “toxic astrocyte” phenotype.

Efficacy of respective drugs was analyzed by astrocyte-driven secretion of C3, a marker used to identify toxic astrocytes in MS lesions (9,11).

Drugs were first tested at 10 μM, but at this concentration, many drugs induced >75% cytotoxic effects on astrocytes (i.e., MTT < 75% of that of the control treatment; Supplementary Table 5). Thus, the entire screen was repeated with 100-fold lower drug concentrations (i.e., 100 nM), which better reflects *in vivo* achievable concentrations of tested drugs in humans (Figure 3).

**Figure 3:**
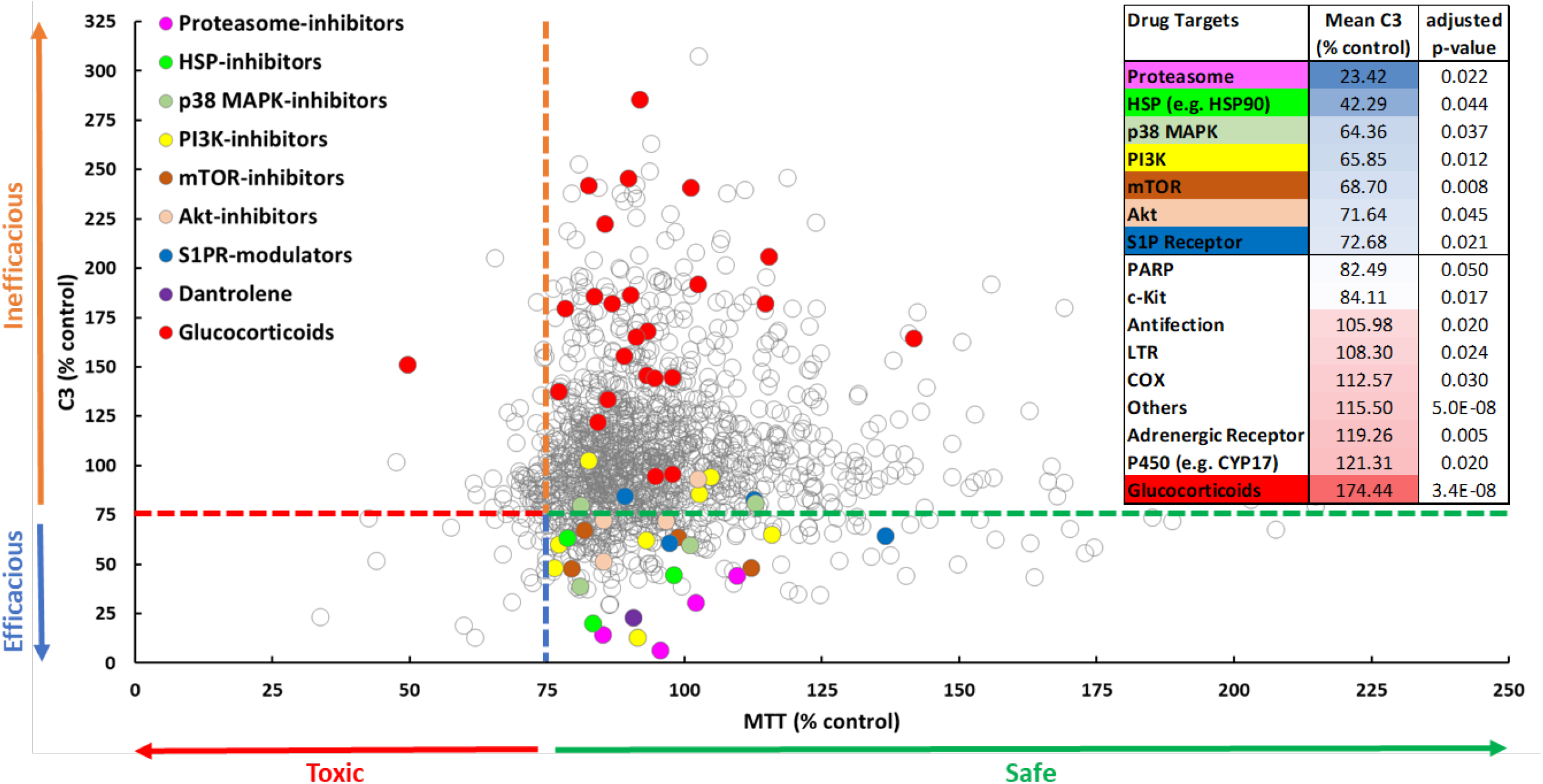
Toxicity (MTT) and efficacy of each drug (at 100 nM) in blocking secretion of C3 was analyzed. Effect of each drug was normalized with respect to control (DMSO) treatment: ([C3 secretion by stimulated astrocytes with a drug treatment – C3 secretion by unstimulated astrocytes] / [C3 secretion by stimulated astrocytes with DMSO treatment – C3 secretion by unstimulated astrocytes]) * 100. 1431 drugs (n = 1) are represented here; significantly efficacious drugs at group level (mean C3 < 75% of control and adjusted p-693 value < 0.05) are represented with respective colors.

### Identification of signaling pathways important in inflammation-induced transformation of astrocytes towards a toxic phenotype

For pathway analysis, drugs with common therapeutic target that were not cytotoxic (i.e., MTT<75% of control) were grouped together. For these groups we calculated mean % inhibition of C3 secretion and formally tested the statistically significant group’s effects. The full results are in Supplementary Table 6, while Figure 3 provides results for relevant drug categories. Inhibitors of multiple pathways that interact together and participate in endoplasmic reticulum (ER) stress response (i.e., unfolder protein response; UPR) reached formal statistical significance in this analysis. Specifically, inhibitors of the following targets (arranged in the order of descending potency) reduced C3 secretion from inflammation-induced astrocytes by at least 25%: 1) proteasome, 2) heat shock protein 90 (HSP90), 3) p38 mitogen-activated protein kinase (p38 MAPK), 4) phosphoinositide 3-kinases (PI3K), 5) mammalian target of rapamycin (mTOR), 6) Akt, and 7) sphingosine 1-phosphate (S1P) receptor. Additionally, dantrolene, an anti-spasticity drug that inhibits calcium release from the ER of CNS cells via inhibiting ryanodine receptor (RyR), also reliably inhibited secretion of C3 with high potency in the drug library screen (i.e., 77.7 % inhibition). Inhibitors of c-kit and poly ADP ribose polymerase (PARP) also achieved statistical significance (FDR-adjusted p <0.05), although their inhibitory effect did not reach 75%. The summary of signaling pathways found inhibitory in our screen and their physiological relationship is depicted in Figure 4.

**Figure 4:**
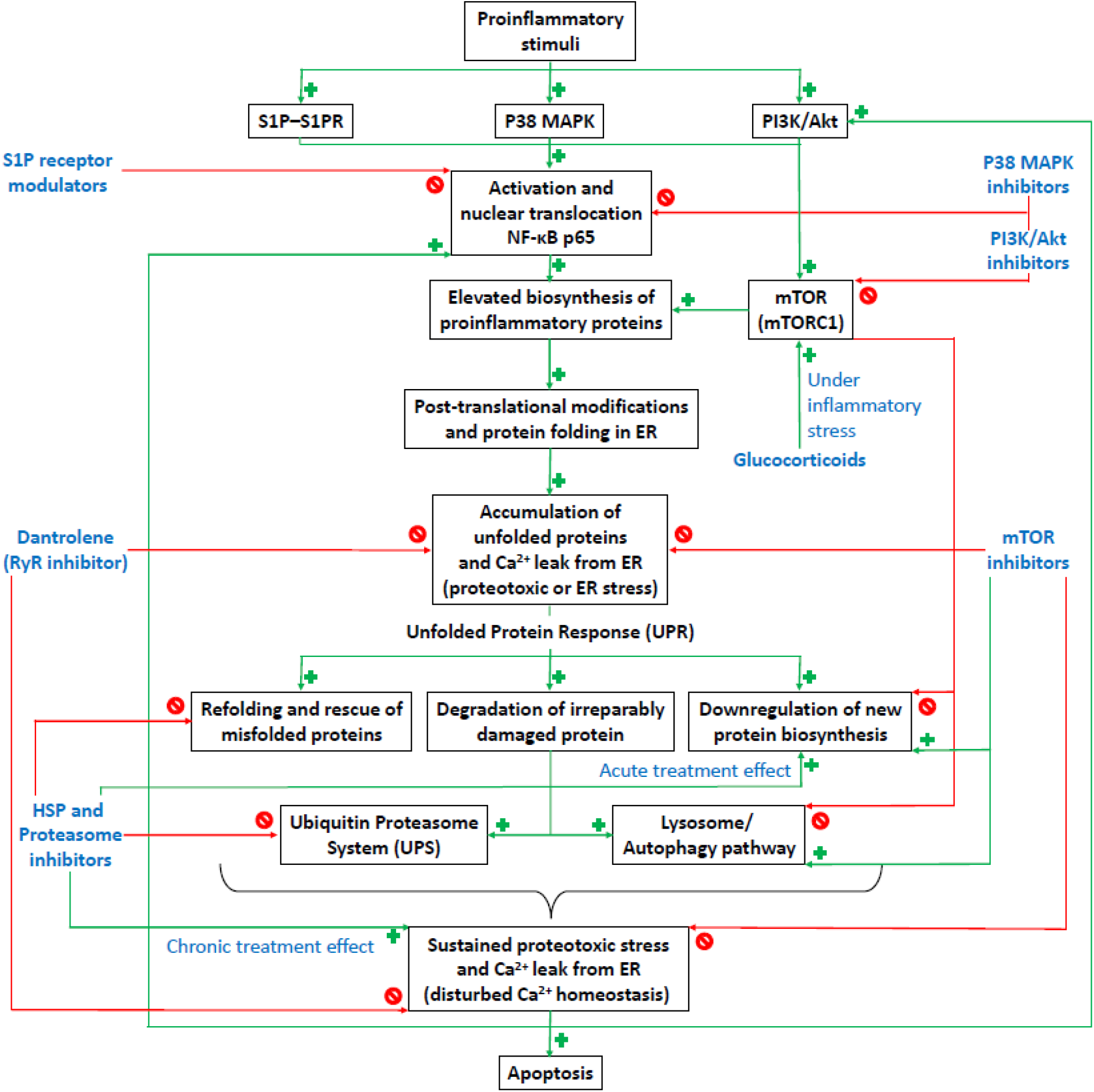
The summary of signaling pathways found inhibitory in our screen and their physiological relationships. Our results suggest that NFκB-p65 mediated transcription of proinflammatory mediators plays a central role in induction of toxic astrogliosis. Abnormally high protein secretion during inflammatory stress may overwhelm the protein folding capacity of the ER, causing accumulation of unfolded proteins, leading to disturbed Ca2+-homeostasis and proteotoxic stress. Sustained ER stress further exacerbates the toxic astrocyte-mediated proinflammatory response and under severe stress, it may lead to cell death. Drugs which can alleviate NFκB-mediated proinflammatory responses and relieve ER stress may be effective in blocking astrocyte-mediated toxicity during MS progression. Green color and ‘+’ sign indicate positive regulation. Red color and ‘stop’ sign indicate negative regulation.

Surprisingly, most glucocorticoids induced secretion of C3 (174.44 % of control) from inflammation-induced astrocytes. Although other drug groups also stimulated C3 secretion from toxic astrocytes with formal statistical significance, the group effects were lower than 125%.

These stimulatory drug targets include cytochrome P450s (CYPs), adrenergic receptor, cyclooxygenase (COX), leukotriene receptor (LTR), and anti-infection agents. Drugs that did not pertain to any specific category and thus were grouped as “others” also achieved a minimal stimulatory effect that reached formal statistical significance (Figure 3).

### Concentration-response curves for selected drugs

Because the drug library screen was performed only in two concentrations (10 μM and 100 nM), the representative drugs from the most effective target categories and other drugs of potential interest were validated in independent experiments, using a concentration curve of 0, 10, 100, and 1,000 nM concentrations. 50% inhibitory concentrations (IC_50_) were calculated from these curves (Figure 5).

**Figure 5:**
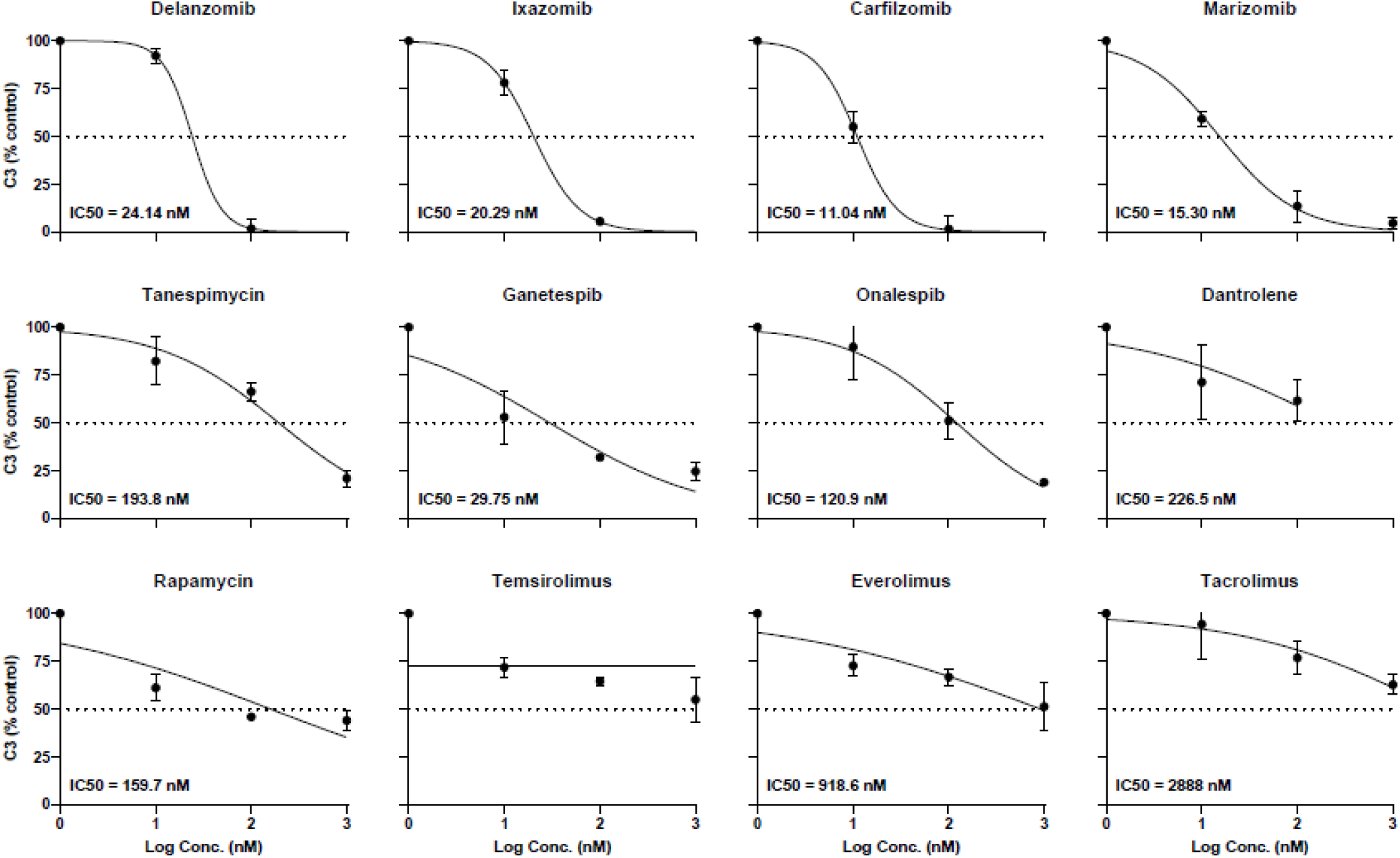
Concentration-response curves for selected drugs. Efficacy of blocking C3 secretion of proteasome-, HSP90-, and mTOR-inhibitors, and dantrolene were tested at 0, 10, 100, and 1000 nM concentrations. Drugs were tested in triplicates for each concentration. Mean ± standard deviations are represented here. The dotted line indicates a 50% reduction in C3 secretion, compared to control, and IC_50_ for each drug are depicted on respective graphs. For better representation, y-axis limits are selected to be from 0 to 100%, but some datapoints and error bars may be out of axis limits.

All tested proteasome inhibitors (Delanzomib, Ixazomib, Carfilzomib and Marizomib) blocked C3 secretion from inflammation-induced astrocytes at low concentrations (IC_50_ < 25 nM). Out of three tested HSP90 inhibitors, Ganetespib had the highest potency (IC_50_ = 29.75 nM), while the other two drugs, Tanespimycin and Onalespib, had an IC_50_ 193.8 and 120.9 nM, respectively. Within tested mTOR-inhibitors, Rapamycin had lowest IC_50_ (159.7 nM), and Everolimus and Tacrolimus had IC_50_ 918.6 and 2888 nM respectively; While, based on tested concentrations, IC_50_ for Temsirolimus cannot be determined. Dantrolene, an RyR-antagonist, had projected IC_50_ 226.5 nM.

### Effects of selected drugs on inflammation-induced astrocyte-mediated neuronal apoptosis

To validate that inhibition of C3 secretion from toxic astrocytes also inhibits neuronal apoptosis, we tested the effects of selected drugs on toxic astrocyte-induced neuronal apoptosis *in vitro*. SK-N-SH cells were treated with astrocyte-conditioned medium (treated with unstimulated or stimulated PBMC supernatants in the presence of a respective drug or DMSO; 50% v/v), and neuronal apoptosis was analyzed 24 hours later. Treatment with supernatants from inflammation-induced astrocytes significantly enhanced neuronal apoptosis compared to supernatants from unstimulated astrocytes (Figure 6). Most tested drugs prevented inflammation-induced astrocyte-mediated neuronal apoptosis, except Delanzomib, Ixazomib and Ganetespib. Delanzomib treatment significantly elevated neuronal apoptosis, suggesting its direct neurotoxic effects.

**Figure 6:**
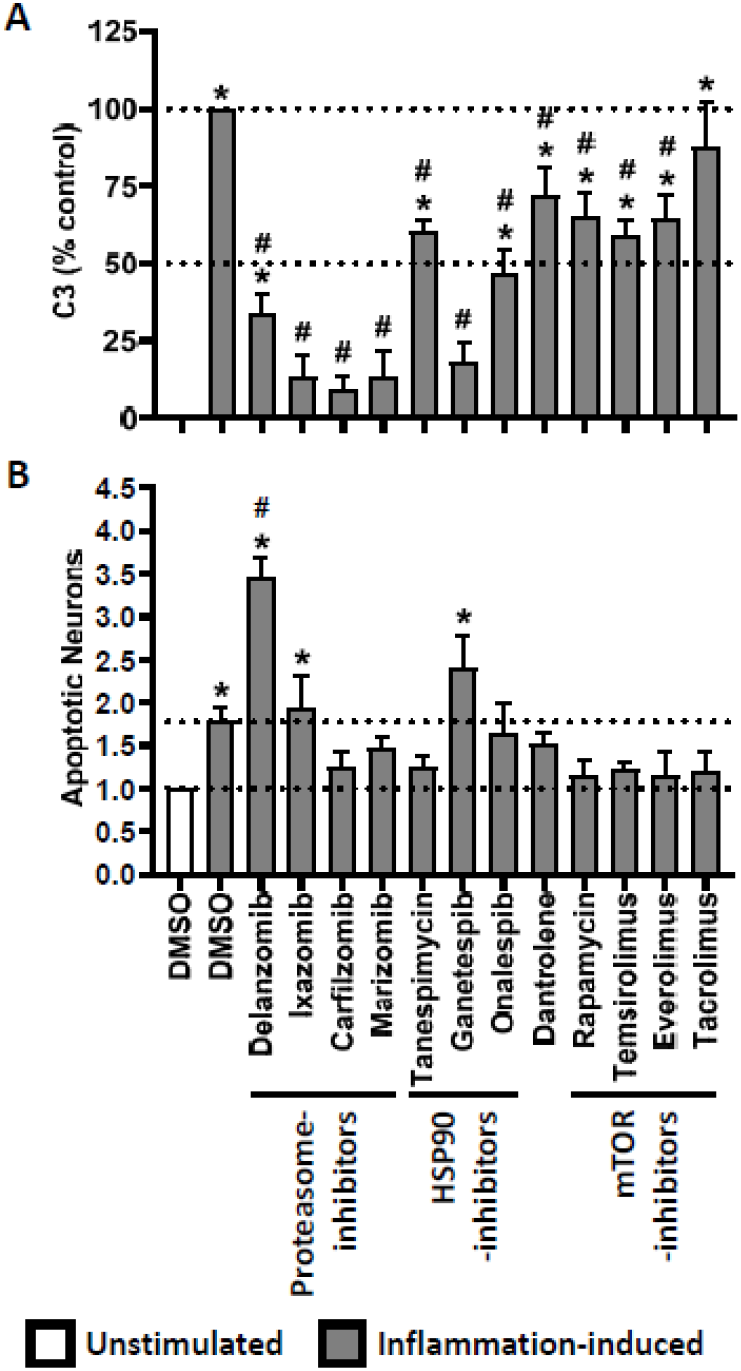
(A) Absolute (24h – 0h) secretion of C3 by astrocytes under different culture conditions were analyzed using ELISA. The effect of each tested drug (100 nM) was normalized with respect to control (DMSO) treatment: ([C3 secretion by inflammation-induced astrocytes with a drug treatment – C3 secretion by unstimulated astrocytes] / [C3 secretion by inflammation-induced astrocytes with DMSO treatment – C3 secretion by unstimulated astrocytes]) * 100. (B) Unstimulated or stimulated ACM-mediated neuronal apoptosis was analyzed using Annexin V immunostaining and flow cytometry; Normalized % of apoptotic (Annexin V+) neurons. C3 secretion and neuronal apoptosis across different culture conditions (n = 3) were compared using ANOVA (Tukey’s multiple comparisons test). *p < 0.05 vs unstimulated + DMSO and #p < 0.05 vs. inflammation-induced + DMSO.

### Proteomic analyses of supernatants from inflammation-induced astrocytes in the presence of selected drugs

While we performed drug library screening using astrocyte-secretion of C3, because this marker has been widely used as evidence of a toxic astrocyte signature in pathology studies (9), C3 is unlikely to be directly neurotoxic. Proteomic analysis identified multiple biomarkers secreted by inflammation-induced astrocytes, out of which only SERPINA3 validated significant relationship to MS severity in an independent validation cohort (Figure 2).

Therefore, to understand the efficacy of drugs that inhibit C3 secretion on the proteome of inflammation-induced astrocytes, we selected three representative drugs (Ganetespib [HSP90-inhibitor], Dantrolene [RyR-antagonist], and Rapamycin [mTOR-inhibitor]) and studied their influence on the secretome of inflammation-induced astrocytes using SOMAscan. Ganetespib was selected based on its reproducibly strong efficacy on C3 secretion, while Dantrolene and Rapamycin were studied for their potential use as candidate inhibitors of toxic astrocytes in proof-of-principle clinical trial in MS.

Ganetespib strongly reduced secretion of all biomarkers previously identified as part of a “toxic astrocyte signature” (Figure 7). However, Ganetespib also inhibited secretion of proteins from astrocytes that were not activated by inflammatory stimuli (Supplementary Table 7). This suggest inhibition of physiological functions of astrocytes, which may have detrimental effects on neurons *in vivo*. Dantrolene reduced secretion of most of the “toxic astrocyte” biomarkers, including SERPINA3. However, it also elevated secretion of MMPs (MMP-10 and 12). In contrast, Rapamycin did not have potent effect on secretion of most of the “toxic astrocyte” biomarkers, except CX3CL1, C3, MMP13 and IL1RL1. Rapamycin elevated secretion of CCL20, TNFAIP6 and CFB.

**Figure 7:**
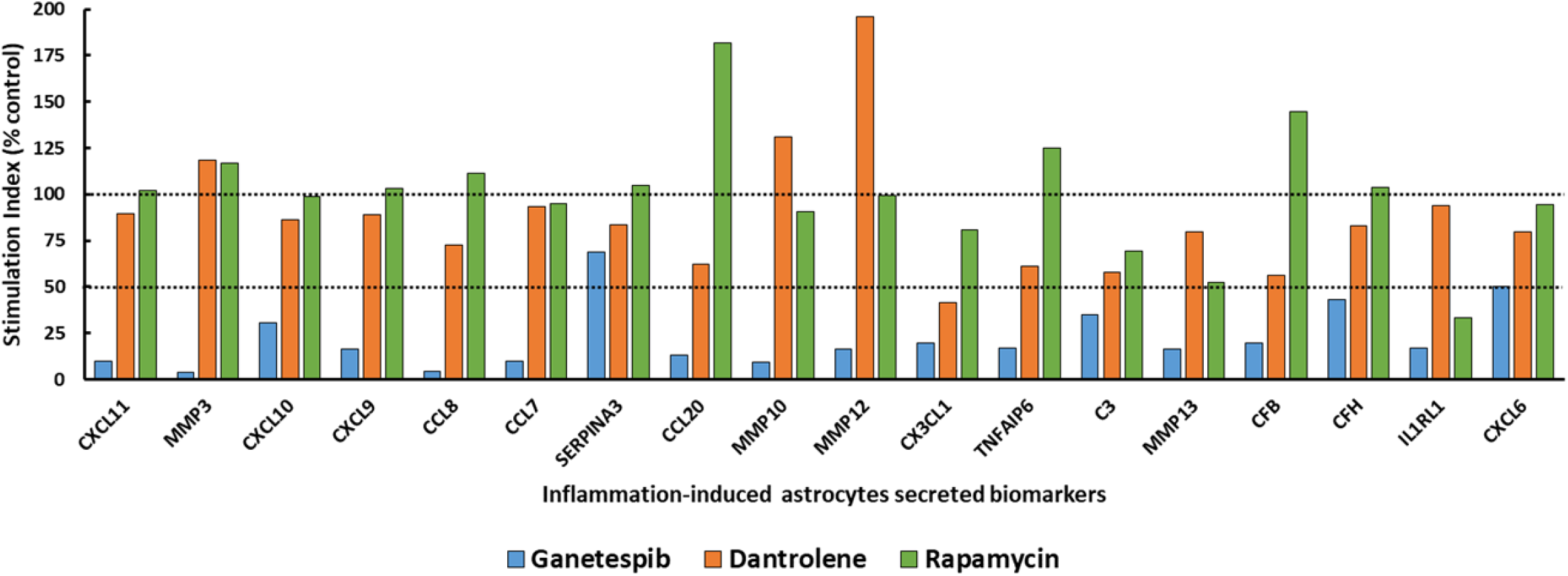
Normalized stimulation indices for inflammation-induced astrocytes secreted biomarkers after treatment with Ganetespib, Dantrolene and Rapamycin. Stimulation index for each protein, under DMSO or respective drug treatment, was calculated by taking the ratio of SOMAscan values under inflammation-induced culture conditions versus unstimulated culture conditions. Stimulation indices for each drug were then normalized with respect to DMSO treatment (stimulation index [% control]).

## Discussion

There are two approaches to drug library screening: *in vitro* assays and *in vivo* experiments in short-lived animal species, such as fruit-flies or zebrafish (23,24). Both have limitations: the non-physiological approach of *in vitro* assays risks the possibility that obtained results that do not reproduce the *in vivo* situation, while animal models suffer from differences in physiological and pathogenic mechanisms between lower species and humans. Nevertheless, drugs identified through *in vitro* drug screens validated therapeutic efficacy in humans (25).

We acknowledge following drawbacks of our study: 1) *In vitro* monoculture might have influenced the astrocyte phenotype, as was previously observed on transcriptome level (26); 2) The short-term induction of toxic astrocytes by inflammatory stimuli may not capture spectrum of toxic astrocytes *in vivo*, where long-term exposure to an inflammatory environment may cause epigenetic changes not reversible by pharmacological manipulations; 3) While we selected inhibition of C3 for drug library screen as C3 is broadly used to identify toxic astrocytes in human brain, including MS lesions (9), the mechanism(s) of neurotoxicity by toxic astrocytes have not been elucidated and are unlikely caused by C3. Therefore, inhibition of C3 does not guarantee inhibition of astrocyte-induced neurotoxicity. Thus, every drug screen requires validation through interventional clinical trial(s) to prove or disprove that the targeted mechanism was truly pathogenic.

Being mindful of the limitations, we employed strategies to maximize the clinical relevance of drug library screen: we analyzed overlap between the toxic astrocyte-secreted proteome in our *in vitro* model and previously published studies and found overlap for CXCLs (11), SERPINA3 (9,11,27), C3 (9), CFB (9) and MMPs (28). This suggests that our assay captures most of the *in vivo* phenotypical change of ‘toxic astrogliosis’.

C3 has been widely used as a marker of the A1/toxic astrocytes (9,29,30) and Liddelow et al found C3 protein mainly expressed in astrocytes (9). However, under neuroinflammatory conditions, CSF C3 concentrations cannot be solely attributed to astrocytes because on an mRNA level, C3 is mainly expressed in microglia and immune cells of myeloid lineage (Supplementary Table 8). This is aligned with our observation that CSF C3 is significantly elevated only in RR-MS. This suggests that during the formation of MS lesions, most C3 either originates from serum and reaches CSF due to blood-brain barrier (BBB) opening or is secreted by cells of the myeloid lineage recruited to acute MS lesions (2,3). Thus, while CSF C3 cannot be a reliable biomarker of toxic astrogliosis *in vivo*, the same problem is not associated with *in vitro* astrocyte monocultures, used in library screen.

From all inflammation-induced astrocyte-secreted proteins, only SERPINA3 CSF levels are astrocyte-specific, as infiltrating immune cell or other CNS cells has minimal SEPINA3 mRNA expression (Supplementary Table 8). The relevance of the CSF SERPINA3 as a biomarker of toxic astrocytes is supported by animal studies. Genomic analysis of astrocytes from pro-inflammatory stimuli-treated mice has shown robust increase in SERPINA3 (11). SERPINA3 also induced toxic effects on cortical murine neuron cultures (27), suggesting its direct pathogenic role. Whether this pathogenicity is true in humans requires clinical trial evidence. Nevertheless, we conclude that CSF SERPINA3 is the best biomarker for *in vivo* pharmacodynamic readout of the inhibitory effect therapies on toxic astrogliosis.

The first important insight from our study identifies small molecules among FDA-approved MS drugs with inhibitory effect on toxic astrocytes. One study reported that *in vitro* pre-treatment of astrocytes with dimethyl fumarate (DMF; 25 µM) reduced secretion of proinflammatory cytokines and oxidative stress following stimulation with IL1β (31). We too observed that DMF inhibited C3 secretion by astrocytes at a non-physiologically high (10 µM) concentration but not at 100 nM. A 25 µM dose is almost 1000-fold higher than the peak measured concentration in human blood (32) and the concentrations in the CNS are likely even lower. This highlights the essential limitation of testing inhibitory effects with non-physiological drug concentrations. In contrast, we based our conclusions on 100 nM drug concentration screen and validated the most promising drugs in dose titration experiments (down to 10 nM).

In contrast to DMF, our drug library screen identified S1P receptor modulators as inhibiting C3 secretion from toxic astrocytes. Indeed, immunostaining of postmortem MS brains showed elevated expression of S1P receptors on astrocytes in MS lesions (33), suggesting a role of S1P receptors in the induction of toxic astrogliosis. Additionally, pretreatment of human induced pluripotent stem cells (iPSCs)-differentiated astrocytes with fingolimod or siponimod reduced secretion of proinflammatory cytokines in the presence of inflammatory stimuli via blocking activation and nuclear translocation of the NFκB-p65 (34). Our results expand on these data and suggest that S1P receptor modulators may also partially block the process of toxic astrogliosis *in vivo*. We plan to test this hypothesis in future studies evaluating the effect of S1P receptor modulators on CSF SERPINA3 levels.

While, glucocorticoids, effective inhibitors of neuro-inflammation (35,36), reproducibly elevated secretion of C3 from inflammation-induced astrocytes *in vitro*, there is currently no experimental evidence that increased secretion of C3 captures the complete neurotoxic phenotype of astrocytes. Thus, it will be important to analyze CSF SERPINA3 levels in patients treated with steroids in future studies.

Next, we’ll provide integrative analysis of signaling pathways that library screen identified as contributing to formation of toxic astrocytes (please refer to Figure 4). Among these, HSP and proteasome inhibitors were the most potent. In response to inflammation, astrocytes initiate protein biosynthesis to amplify the inflammatory response. Most of these synthesized proteins are secreted, requiring posttranslational modifications and proper folding in the ER before entering the Golgi apparatus. High protein secretion may overwhelm protein folding capacity, resulting in accumulation of unfolded proteins, known as proteotoxic or ER stress. This triggers UPR response aimed to restore homeostasis, consisting of the following actions/pathways: 1) refolding of misfolded proteins, 2) degradation of irreparably damaged proteins via the ubiquitin proteasome system (UPS) and lysosome/autophagy-mediated pathways, and 3) downregulation of new protein biosynthesis (37,38). The UPR is initially protective, but under sustained stress, it induces further inflammation and eventually triggers apoptosis (39).

By inhibiting protein folding and degradation of misfolded proteins, HSP and proteasome inhibitors exacerbate ER stress and stall protein synthesis. Indeed, Ganetespib, an HSP-inhibitor, reduced secretion of all astrocyte-secreted proteins, both physiological and inflammation-induced. Clearly, such a strategy is not sustainable long-term, as it will likely cease essential astrocyte-mediated functions that may not be consistent with the survival. Even in oncology, these drugs can be administered only short-term. Thus, we excluded these drugs/pathways from consideration for treatment of neurodegenerative diseases.

Another class of drugs identified through our drug library screening are PI3K/Akt/mTOR-inhibitors. This pathway is known to positively regulate the proinflammatory response via upregulating activation and nuclear translocation of NFκB-p65 (40,41). Particularly mTOR signaling plays a vital role in astrocytic proliferation and production of proinflammatory mediators during stress (42). mTOR has a bidirectional crosstalk with ER stress: 1) mTOR signaling works upstream and exacerbates ER stress via upregulating protein biosynthesis and downregulating clearance of damaged proteins through the lysosome/autophagy-mediated pathways, and 2) mTOR signaling also works downstream of ER stress, where sustained ER stress activates mTOR signaling, leading to inflammation and cell death (41,43,44). Though our results showed that PI3K/Akt/mTOR-inhibitors inhibit secretion of C3 from inflammation-induced astrocytes, Rapamycin failed to inhibit secretion of SERPINA3, and actually increased secretion of some pro-inflammatory molecules, such as CCL20, CCL8. This is an intriguing observation as it suggests that different pathways mediate secretion of C3 versus other markers of toxic astrocytes, like SERPINA3. Because the reviewed literature, supported by observed correlation of CSF SERPINA3 with MS severity assigns a stronger role to SERPINA3 than C3 as the marker of pathogenic astrocytes, PI3K/Akt/mTOR inhibitors may not inhibit the most relevant neurotoxic functions.

Dantrolene, an RyR-antagonist, significantly and reproducibly reduced secretion of C3, but also SERPINA3, CCL20, CCL8 from inflammation-induced astrocytes. Under normal conditions, the ER has at least a three-fold higher Ca^2+^ concentration than that of the cytosol, which is crucial for normal protein folding. Early ER stress dysregulates RyR functioning (45), causing Ca^2+^ leak from the ER, which further disturbs normal protein folding. During sustained ER stress, dysregulated RyR-mediated Ca^2+^ leak from the ER positively regulates UPR-mediated inflammation and cell death (45,46), suggesting that dantrolene should alleviate ER stress. This would mean that drugs with opposing mechanisms on ER stress (i.e., HSP/proteasome inhibitor exacerbating and dantrolene alleviating ER stress) are both efficacious in suppressing generation of toxic astrocytes. How can we resolve this discrepancy? One possibility is that the neurotoxic astrocytes are not induced directly by inflammatory stimuli, which signal via PI3K/Akt/mTOR and p38MAPK. Instead, the result of this pro-inflammatory signaling is robust protein synthesis and ER stress. Perhaps it is the failing ER stress phase of the astrocytic response to inflammation that generates their true neurotoxic phenotype. The HSP/proteasome inhibition tips the astrocyte one way (shutting off protein synthesis completely and eventually causing cell death), while dantrolene blocks the pathogenic transformation of astrocytes by blocking Ca^2+^ release from the failing ER to cytosol and/or to mitochondria. Clearly, this hypothesis will need to be tested in future studies. Nevertheless, since dantrolene is FDA-approved for treatment of spasticity and has been applied in this indication to MS patients long-term, it is an immediately available candidate for testing in proof-of-principle clinical trials its efficacy on toxic astrocyte inhibition. Its major drawback is serious hepatotoxicity with doses over 400 mg/day (47).

In conclusion, this study elucidated signaling pathways associated with transformation of astrocytes to a toxic phenotype and identified candidate drugs for clinical testing. The first step in proving efficacy of any of these agents should be demonstrating their pharmacodynamic effect on CSF SERPINA3 levels, which correlate with MS severity.

## Data Availability

The authors confirm that the data supporting the findings of this study are available within the article and its supplementary materials.

## Acknowledgments

We thank Elena Romm for processing of CSF samples. We thank Dr. Alison Wichman and research nurses Mary Sandford and Tiffany Hauser for expert patient care and patient care coordinator Michelle Woodland for patient scheduling. Finally, we thank all the patients, their caregivers and healthy volunteers, without whom this work could not be possible.

**Abbreviations**
MS: multiple sclerosis
CNS: central nervous system
RRMS: relapsing-remitting MS
PMS: progressive MS
CSF: cerebrospinal fluid
FDA: Food and Drug Administration
PBMCs: peripheral blood mononuclear cells
PBS: phosphate-buffered saline
LPS: lipopolysaccharide
TNFα: tumor necrosis factor α
IL1α: interleukin 1 α
C3: complement component 3
SERPINA3: serine protease inhibitor family A member 3
EDSS: Expanded Disability Status Scale
CombiWISE: Combinatorial Weight-Adjusted Disability Scale
MRI: magnetic resonance imaging
COMRIS-CTD: Composite MRI scale of CNS tissue destruction
MSSS: Multiple Sclerosis Severity Score
ARMSS: Age Related Multiple Sclerosis Severity
MSDSS: Multiple Sclerosis Disease Severity Scale
SP-MS: secondary progressive MS
PP-MS: primary progressive MS
HD: healthy donors
SOMAscan: Slow Off-rate Modified Aptamers scan
DMSO: dimethyl sulfoxide
MTT: 3-(4,5-dimethylthiazol-2-yl)-2,5-diphenyltetrazolium bromide
ANOVA: Analysis of Variance
FDR: False Discovery Rate
CXCL: C-X-C motif chemokine ligand
MMP: matrix metalloproteinases
CCL: C-C motif chemokine ligand
CX3CL1: C-X3-C motif chemokine ligand 1
TNFAIP6: TNFα induced protein 6
CF: complement factors
IL1RL1: interleukin 1 receptor like 1
ER: endoplasmic reticulum
UPR: unfolder protein response
HSP90: heat shock protein 90
p38 MAPK: p38 mitogen-activated protein kinase
PI3K: phosphoinositide 3-kinases
mTOR: mammalian target of rapamycin
S1P: sphingosine 1-phosphate
RyR: ryanodine receptor
PARP: poly ADP ribose polymerase
CYPs: cytochrome P450s
COX: cyclooxygenase
LTR: leukotriene receptor
IC_50_: 50% inhibitory concentrations
BBB: blood-brain barrier
GSK-3β: glycogen synthase kinase-3β
TMT: trimethyltin chloride
DMF: dimethyl fumarate
iPSCs: induced pluripotent stem cell
UPS: ubiquitin proteasome system

## Supplementary Table Legends

**Supplementary Table 1:** Cohort (training- and validation-cohort), diagnosis (HD, RRMS, SPMS and PPMS), clinical outcome measures (EDSS, CombiWISE, COMRIS-CTD, MSSS, ARMSS and MSDSS) and age- and gender-adjusted CSF SOMAscan (1.1K platform; only toxic astrocyte-specific markers, 17 proteins) data of all subjects (n = 333). Patients were recoded and personal identification information (PII, such as age, gender and clinic visit dates) were excluded.

**Supplementary Table 2:** SOMAscan (1.3K platform, 1317 proteins) data of astrocyte culture supernatants, under different culture conditions (unstimulated and stimulated), at 0 and 24 hours. Stimulation index (stimulated [24h / 0h] / unstimulated [24h /0h]) for each protein were calculated.

**Supplementary Table 3:** Within training cohort, differences for inflammation-induced, astrocytes-secreted biomarkers’ SOMAscan values (age- and gender-adjusted) across disease diagnostic subgroups (HD, RR-MS and P-MS) were analyzed using Kruskal-Wallis ANOVA (Dunn’s multiple comparisons test). Only statistically significant (adjusted p-value < 0.05) differences were then validated in an independent validation cohort.

**Supplementary Table 4:** Correlations between age- and gender-adjusted SOMAscan values for inflammation-induced, astrocytes-secreted biomarkers and clinical outcome measures were analyzed using Spearman analysis. Only statistically significant (adjusted p-value < 0.05) correlations were then validated in an independent validation cohort. Table represents Spearman’s rank correlation coefficient (ρ) and P values.

**Supplementary Table 5:** Drug library (Selleckchem; FDA-approved Drug Library) screening data. All drugs (1431) were first screened at 10 μM, and then at 100 nM. Toxicity (MTT assay) and efficacy (inhibiting absolute secretion of C3) of each drug were analyzed. MTT and C3 assay results for each plate were normalized based on respective control treatment (stimulated + DMSO, % control). To summarize overall effect of each drug at two different concentrations, mean and standard deviation (SD) were calculated.

**Supplementary Table 6:** Drugs with common therapeutic targets were grouped together (1431 drugs were grouped into 151 groups). At 100 nM concentration, only for safe drugs (MTT > 75% of control), mean C3 concentrations (% of control) for each group were calculated and compared with control (DMSO; C3 = 100%), p-values were adjusted for multiple comparisons.

**Supplementary Table 7:** SOMAscan (1.3K platform, 1317 proteins) data of astrocyte culture supernatants, under different culture conditions: unstimulated and inflammation-induced, in presence of DMSO or selected drugs (Ganetespib, Dantrolene and Rapamycin; 100 nM), 0 and 24 hours. Stimulation indices for each protein, under DMSO or respective drug treatment, were calculated. Then stimulation indices for each drug were normalized with respect to DMSO treatment (% control).

**Supplementary Table 8:** Relative expression (RNA-Seq analyses) of inflammation-induced, astrocytes-secreted biomarkers, within human CNS and blood cells. Data from publicly available databases were extracted and compiled. Human CNS cells database: https://www.brainrnaseq.org/; Human blood cells database: https://www.proteinatlas.org/about/download.

